# Proteomics screening post pediatric allogeneic hematopoietic stem cell transplantation reveals an association between increased expression of inhibitory receptor FCRL6 on γδ T cells and CMV reactivation

**DOI:** 10.1101/2023.11.02.23297952

**Authors:** Adam Alexandersson, Mikko S Venäläinen, Nelli Heikkilä, Xiaobo Huang, Mervi Taskinen, Pasi Huttunen, Laura L Elo, Minna Koskenvuo, Eliisa Kekäläinen

## Abstract

**Objective:** To study kinetics and associations between inflammation related proteins in circulation after pediatric allogenic hematopoietic stem cell transplantation (HSCT) to reveal proteomic signatures or individual soluble proteins associated with specific complications post HSCT.

**Methods:** We used a proteomics method called Proximity Extension Assay to repeatedly measure 180 different proteins together with clinical variables, cellular immune reconstitution, and blood viral copy numbers in 27 children aged 1-18 years during a two-year follow up after allogenic HSCT. Protein profile analysis was done using unsupervised hierarchical clustering and a regression-based method, while Bonferroni-corrected Mann-Whitney U test was used for time point specific comparison of individual proteins against outcome.

**Results:** At 6 months after allogenic HSCT, we could identify a protein profile pattern associated with occurrence of the complications chronic graft-versus-host disease, viral infections, relapse, and death. When protein markers were analyzed separately, the plasma concentration of the inhibitory and cytotoxic T cell surface protein FCRL6 (Fc receptor-like 6) was higher in patients with CMV viremia (log2-fold change 1.5 (p0.00099), 2.5 (p=0.00035) and 2.2 (p=0.045) at time points 6, 12 and 24 months). Flow cytometry confirmed that FCRL6 expression was higher in innate-like γδ T cells, indicating that these cells have a role in controlling CMV reactivation in HSCT recipients.

**Conclusions:** The potentially druggable FCRL6 receptor on cytotoxic T cells appears to have a role in controlling CMV viremia post-HSCT. Our results suggest that system level analysis is a useful addition to the studying of single biomarkers in allogeneic HSCT.

## Introduction

Allogenic hematopoietic stem cell transplantation (HSCT) has the potential to cure several malignant and benign conditions of the bone marrow, but is burdened by risk of complications(1). Routinely used clinical parameters and laboratory tests are insufficient to predict the occurrence of potentially lethal and debilitating complications such as graft-versus-host disease (GVHD)(2) and bacterial, fungal and viral infections(3–5). Reactivation of latent cytomegalovirus (CMV) is one of the main complications after allo-HSCT(6). Tools to predict complications are needed since for example antivirals used to treat CMV reactivations are potentially toxic, and their use should be limited only to patients that clearly benefit from them. Adoptive transfer of pathogen-specific allogenic lymphocytes is still mostly an experimental treatment or prophylaxis option for reactivation of latent viruses; the optimal cellular composition of the adoptive transfer to balance the risk for GvHD and effective antiviral clearance remains unknown(7–9).

Many studies have focused on predicting CMV reactivation but validated biomarkers are still lacking(10). Delayed immune reconstitution is a risk for CMV reactivations(10). CMV-specific CD8+ T-cell cytokine signatures can predict the risk of CMV reactivation, but testing of cellular responses is complicated and not suitable for routine clinical laboratories(11). Testing for individual biomarkers or a panel of soluble plasma biomarkers to predict complications after HSCT would better suit clinical practice. Proteomics approaches after HSCT and results are promising regarding understanding of pathogenesis as well as predicting relapse, non-relapse mortality, acute GVHD (aGVHD) and chronic GVHD (cGVHD), including therapy response(2,12,13). No biomarker or panel of biomarkers is, however, yet widely in clinical use(2).

A benefit of the proteomics and other systems immunology approaches is their hypothesis generating attribute, as opposed to testing a priori formulated research questions(14). To our knowledge, no proteomic studies have been performed prospectively on a well-defined cohort of children receiving HSCT with myeloablative conditioning. In order to better predict the onset and to understand mechanisms of severe complications after HSCT in children, we performed a longitudinal deep analysis on the plasma proteome in relation to clinically relevant outcomes like cGVHD, viral infections, relapse and death.

## Materials and methods

### Patient cohort, sample collection and clinical outcomes

This single-center study from the New Children’s Hospital, Helsinki University Hospital, Finland, followed 27 children with hematological cancer aged 1-18 years for two years after HSCT with myeloablative conditioning. We have earlier published the cohort’s clinical characteristics, including timing of complications(15). In addition to registering clinical disease- and transplant-related features, symptoms and signs, we collected blood samples at time points 3, 6, 12 and 24 months post HSCT. Plasma was isolated and stored frozen. Peripheral blood mononuclear cell (PBMC) isolation was done with gradient centrifugation (Ficoll-Paque PLUS, Cytiva, MA, the USA) and cells were frozen in 10% DMSO in fetal-calf serum and stored frozen in liquid nitrogen or −140°C. We measured viral copy numbers of CMV, human herpesvirus 6 (HHV6), BK virus (BKV) and Epstein-Barr Virus (EBV) at 3, 6, 12 and 24 months, and additionally extracted levels of the same viruses from the patient records, when they had been requested by the clinician. Immune reconstitution (IR) was followed with clinical lymphocyte subtype testing (total CD3+ T cells, CD4+ T helper cells, CD8+ cytotoxic T cells, CD19+ B cells and NK cells). Lymphocytes and subclasses were quantified using accredited flow cytometry for clinical use and viral levels with real-time PCR at an accredited clinical laboratory (HUS Diagnostic Center, Helsinki, Finland). Quality control and descriptive statistics are found in the supplementary material.

Since the spectrum of HSCT complications is diverse and numerous outcome subgroups strain statistical analyses, we created a composite outcome called “complication free”. Complication-free status was attributed to patients who did not suffer any of the following: death during follow-up, relapse during follow-up, cGVHD as per the 2014 NIH criteria(16), aGVHD grade 3 or 4(17), CMV viremia, BKV viremia over 10 000 IU/ml, HHV6 viremia in combination with BKV, or EBV viremia. “Complication-prone” status was correspondingly designated if any of the above findings were present during the follow-up.

The study was approved by the Ethics Committee of the Helsinki University Hospital (HUS/2306/2016), and written informed consent was obtained from all parents and patients above 6 years of age. Baseline demographics are presented in Table 1.

**Table 1.**
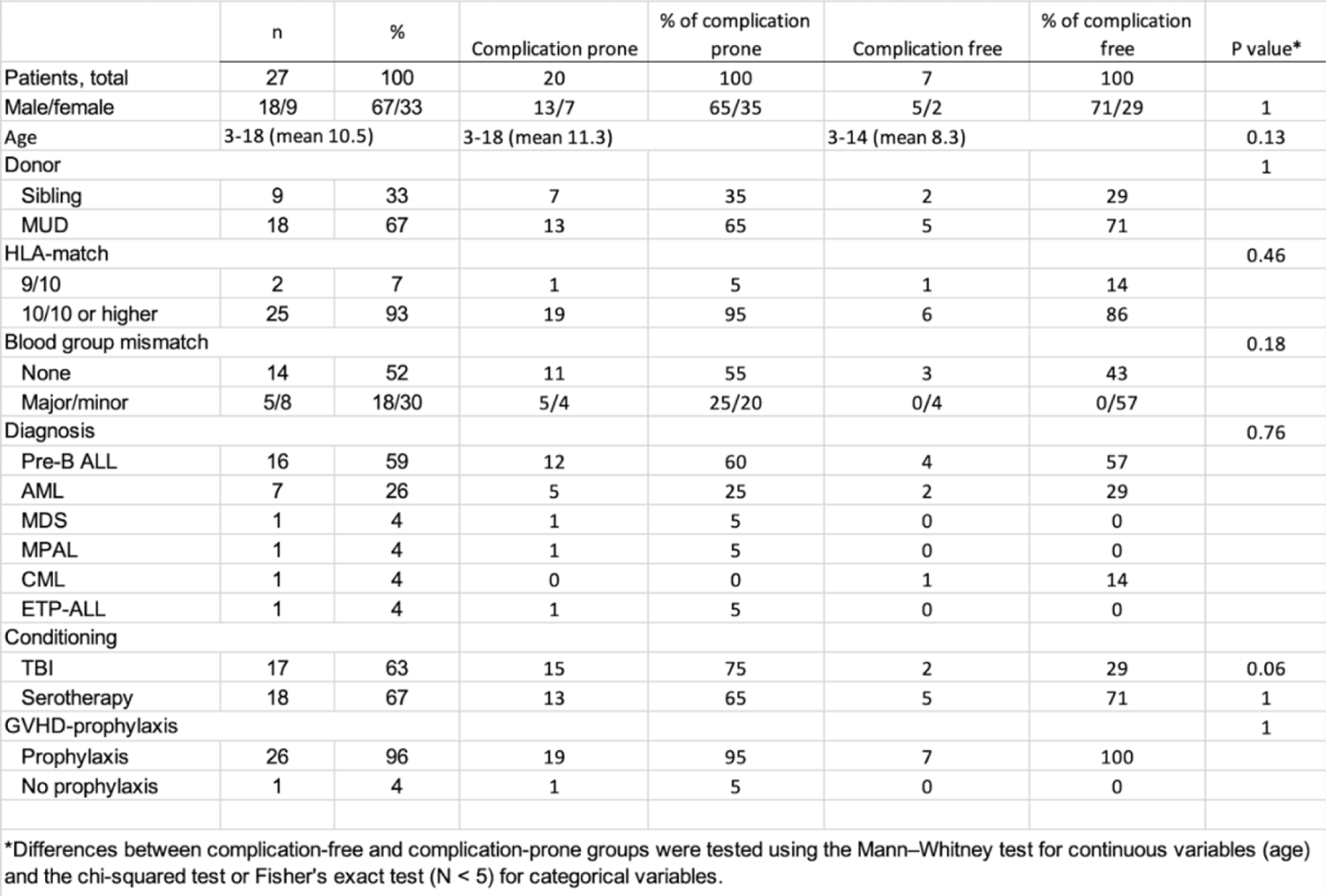
Cohort demographics. MUD, matched unrelated donor; ALL, acute lymphoblastic leukemia; CML, chronic myeloid leukemia; NHL, non-Hodgkin lymphoma; AML, acute myeloid leukemia; MDS, myelodysplastic syndrome; MPAL, mixed phenotype acute leukemia; TBI, total body irradiation; GVHD, graft-versus-host disease. Serotherapy: Alemtuzumab or anti-thymocyte globulin. *Differences between complication-free and complication-prone groups were tested using the Mann–Whitney test for continuous variables (age) and the chi-squared test or Fisher’s exact test (N < 5) for categorical variables.

|  | n | % | Complication prone | % of complication prone | Complication free | % of complication free | P value* |
| --- | --- | --- | --- | --- | --- | --- | --- |
| Patients, total | 27 | 100 | 20 | 100 | 7 | 100 |  |
| Male/female | 18/9 | 67/33 | 13/7 | 65/35 | 5/2 | 71/29 | 1 |
| Age | 3-18 (mean 10.5) |  | 3-18 (mean 11.3) |  | 3-14 (mean 8.3) |  | 0.13 |
| Donor |  |  |  |  |  |  | 1 |
| Sibling | 9 | 33 | 7 | 35 | 2 | 29 |  |
| MUD | 18 | 67 | 13 | 65 | 5 | 71 |  |
| HLA-match |  |  |  |  |  |  | 0.46 |
| 9/10 | 2 | 7 | 1 | 5 | 1 | 14 |  |
| 10/10 or higher | 25 | 93 | 19 | 95 | 6 | 86 |  |
| Blood group mismatch |  |  |  |  |  |  | 0.18 |
| None | 14 | 52 | 11 | 55 | 3 | 43 |  |
| Major/minor | 5/8 | 18/30 | 5/4 | 25/20 | 0/4 | 0/57 |  |
| Diagnosis |  |  |  |  |  |  | 0.76 |
| Pre-B ALL | 16 | 59 | 12 | 60 | 4 | 57 |  |
| AML | 7 | 26 | 5 | 25 | 2 | 29 |  |
| MDS | 1 | 4 | 1 | 5 | 0 | 0 |  |
| MPAL | 1 | 4 | 1 | 5 | 0 | 0 |  |
| CML | 1 | 4 | 0 | 0 | 1 | 14 |  |
| ETP-ALL | 1 | 4 | 1 | 5 | 0 | 0 |  |
| Conditioning |  |  |  |  |  |  |  |
| TBI | 17 | 63 | 15 | 75 | 2 | 29 | 0.06 |
| Serotherapy | 18 | 67 | 13 | 65 | 5 | 71 | 1 |
| GVHD-prophylaxis |  |  |  |  |  |  | 1 |
| Prophylaxis | 26 | 96 | 19 | 95 | 7 | 100 |  |
| No prophylaxis | 1 | 4 | 1 | 5 | 0 | 0 |  |

### Proteome analysis

For proteome analysis we used the Proximity Extension Assay (Olink, Uppsala, Sweden) on freshly thawed frozen plasma samples at time points 3, 6, 12, and 24 months post-HSCT. In short, Proximity Extension Assay is a proteomics method where two target-specific oligo-conjugated antibodies bind their respective epitopes that are very closely situated on the target, allowing the oligos to hybridize. After this, only the hybridized oligonucleotide sequences are detected with qPCR. The method is highly specific and enables a relative quantification of large amounts of protein targets simultaneously in a longitudinal sample series(18). We used two Olink panels (“Inflammation” and “Immune response”) consisting of 92 proteins each (supplementary material, Table S1). The Olink analysis service was provided by the Biomedicum Functional Genomics Unit at the Helsinki Institute of Life Science and Biocenter Finland at the University of Helsinki.

### Flow cytometry

For FCRL6 flow cytometric analysis, PBMCs from 17 patients collected at 3, 6, 12, and 24 months post-transplantation were used as available (supplementary material, Table S2). Frozen aliquots of PBMCs were thawed in +37°C water bath and transferred to 50 mL conical tubes. Then, 10 mL of +37°C thawing solution (RPMI supplemented with 10% CTL-WASH (Cellular Technology Limited) and 10 ug/mL DNase I) was added slowly, the tube was centrifuged at 350g, 10min in room temperature, and the supernatant was discarded. Cells were washed again with 10mL of thawing solution at 350g for 10 min, adjusting the temperature to +4°C during the centrifugation and resuspended in +4°C staining buffer (PBS supplemented with 10% FCS and 10mM EDTA).

For each individual, 1-2 million cells were washed with 500 ul of staining buffer. The fluorescent-labeled anti-human antibodies are listed in Table S3. The optimal staining concentrations for antibodies were titrated with live PBMCs. Cells were incubated with a mix of antibodies and 1:1000 LIVE/DEAD Fixable Green Dead Cell Stain (Thermo Fisher) diluted in Brilliant Stain Buffer (BD Biosciences) in a final volume of 50 ul for 30 min in +4°C. Cells were washed with 1 mL of staining buffer and run on LSR II Fortessa (BD Biosciences). A median of 78 000 (range 8 000-990 000) viable lymphocytes were registered per each run. Flow cytometric data was analyzed using FlowJo software (version 10.8, BD, Biosciences) using fluorescence-minus-one staining for FCRL6 expression, and biological negative controls when applicable. The full gating strategy is displayed in the supplementary material, Figure S1.

### Statistical methods

Protein concentrations were expressed as Normalized Protein eXpression (NPX) values, an arbitrary unit on log2-scale. Values below qPCR limit of detection (LOD) were treated both by using actual obtained data and, due to the risk of bias and the risk of missing out on relevant associations, by setting values below LOD to missing. However, proteins with over 50% of measured values below LOD were excluded completely from further analyses. The multidimensional protein data was visualized by applying principal component analysis (PCA, supplementary material, Fig. S2) on data at each individual time point. Time-course trends of individual proteins were visualized using sparkline plots generated by applying Loess regression smoothing for complication-free and complication-prone groups.

Unsupervised hierarchical clustering of protein levels was performed using the Euclidean distance and Ward’s method independently at each time point. Prior to clustering, the NPX values were scaled using z-score normalization. Differences in outcome rates between identified clusters were compared using the chi-squared or Fisher’s exact test (n<5). Differences in trends of immune reconstitution between the clusters over time were analyzed using linear mixed effect models with cluster, time and cluster×time interaction term as fixed effects and including a random intercept for patients.

Global correlation matrices were computed and prepared for visualization using R package DGCA (https://pubmed.ncbi.nlm.nih.gov/27846853/). For each time point, the top ten protein pairs with the greatest absolute difference in correlation between complication -free and complication-prone patients were reported. Differentially correlating protein pairs were identified using R package DiffCorr (https://pubmed.ncbi.nlm.nih.gov/23246976/).

Differentially expressed markers for outcomes of interest were identified using maSigPro, a regression-based method designed for identifying differential expression profiles between experimental groups in time-course experiments(19). Here, up to third degree polynomial models were considered and the obtained p values were corrected for multiple comparisons by applying the Bonferroni correction. For each model, we used R²=0.5 as a threshold for acceptance into the group of significant proteins. Additionally, differential expression for outcomes of interest at each individual time point was tested using Bonferroni-corrected Mann-Whitney U test.

All statistical analyses and modeling were carried out using the R statistical computing environment version 4.0.3 (R Core Team, 2016. R: A language and environment for statistical computing. R Foundation for Statistical Computing, Vienna, Austria. URL https://www.R-project.org/). The level of significance was set at p<0.05.

## Results

### Unsupervised clustering of plasma proteome identifies patients with HSCT-related complications and poor immune reconstitution

As depicted in the sparkline plots (Fig. 1), individual protein levels plotted over time showed that almost all measured proteins changed over time with varying patterns in complication-free and complication-prone individuals. Therefore, we decided to first investigate a possible association between a global plasma protein profile and clinical outcome measures, by performing an unsupervised clustering. A heatmap (Fig. 2A), created using hierarchical clustering based on protein levels at 6 months post-HSCT, revealed two clusters with distinctly different outcome profiles. Similar but less clearly separating pattern for two main clusters was visible in other timepoints (supplementary material, Fig. S4). We chose to focus on the 6 month timepoint in the following analyses, since by this time most of the complications had occurred, but the reconstitution of the immune system would not yet be finalized. At this point, all aGVHD had occurred, every viral infection was diagnosed(15) and most cases of cGVHD had begun. Some cases of death and relapse had yet to occur, but all patient-specific complications were registered and included in analysis independent of time of onset, since relevant immunological patterns might be seen before, during, and after debut.

**Fig. 1.**
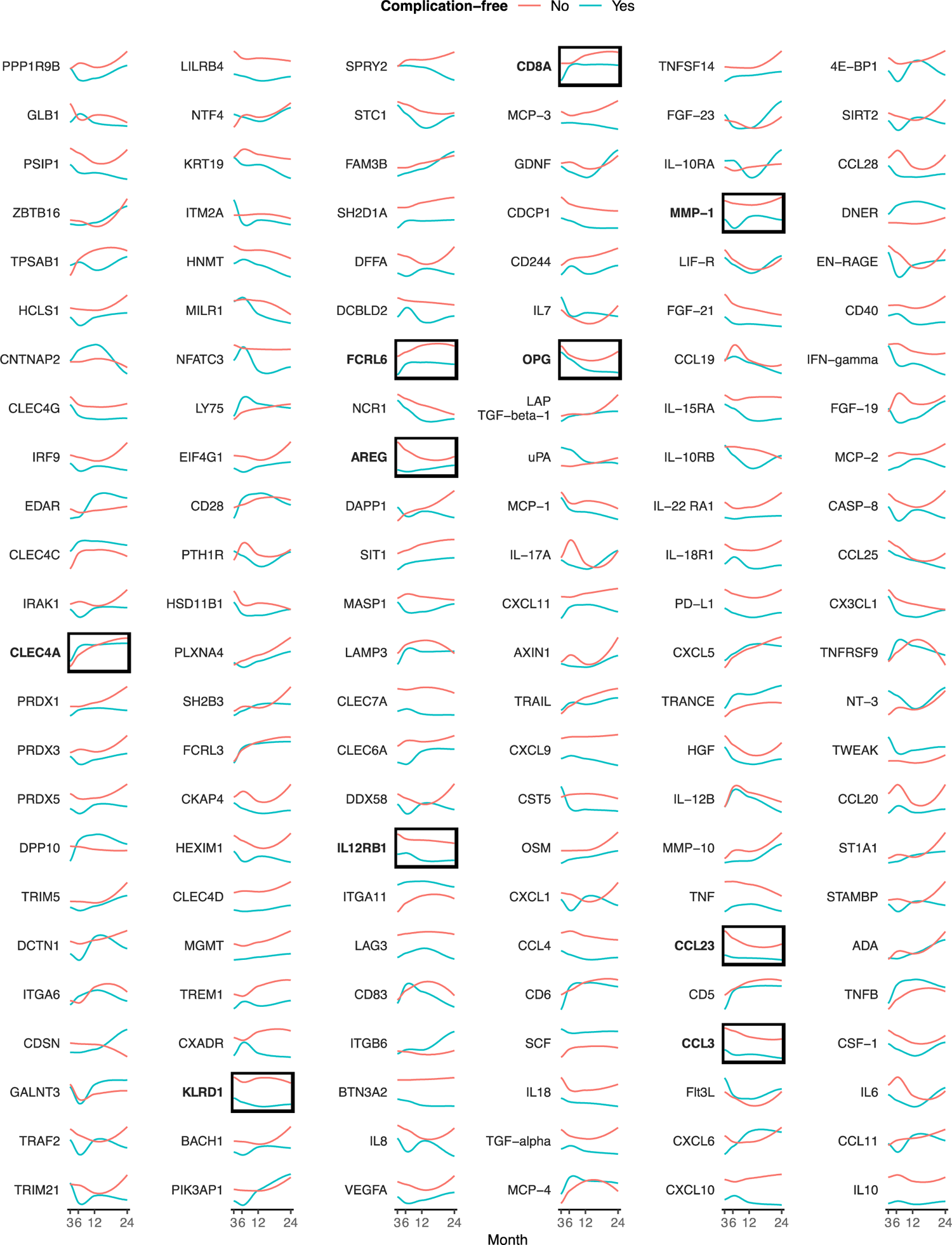
Trajectories of individual proteins. Spark-line plot of protein kinetics over time according to patient grouping by complication-free status (see methods for definition). Highlighted proteins were found to be clearly associated with complications.

**Fig. 2.**
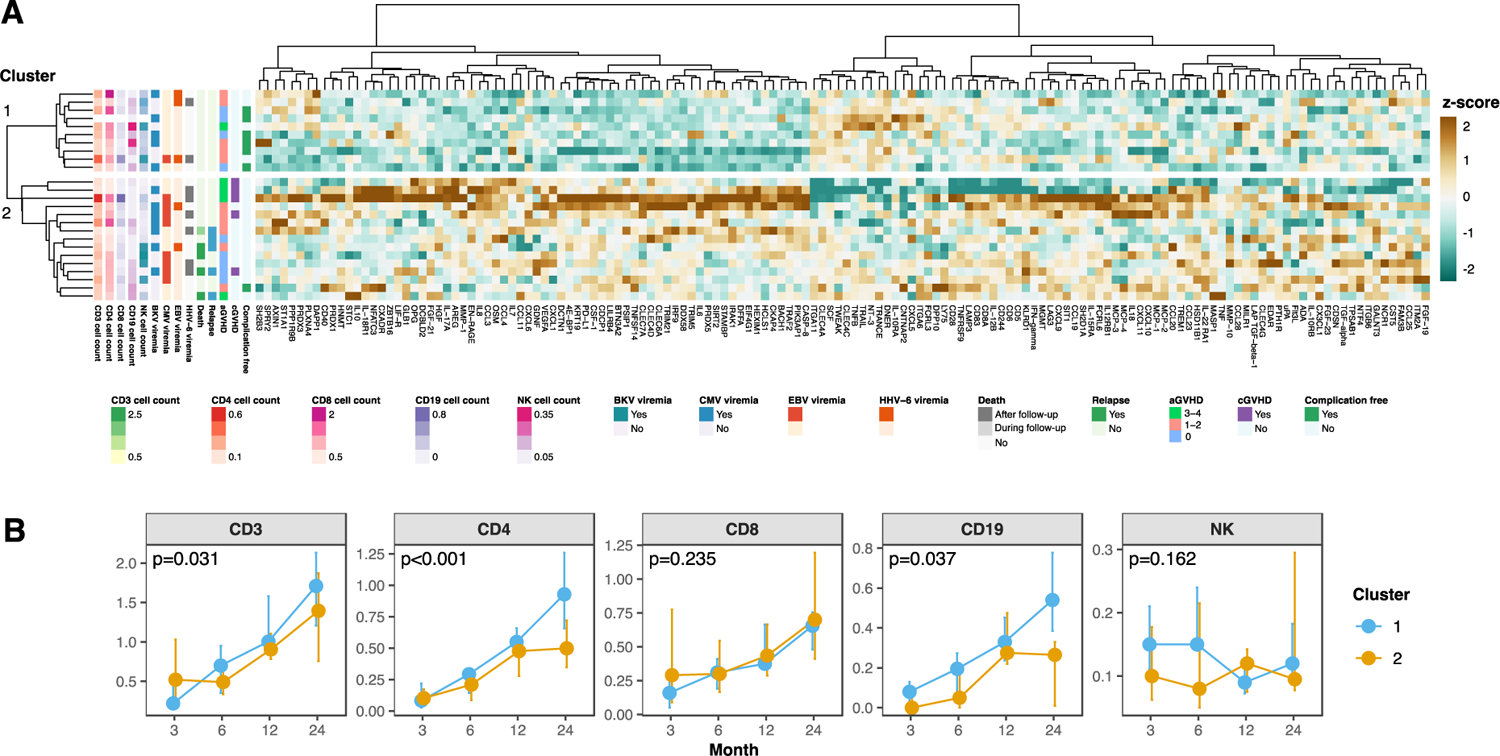
Protein profile in relation to outcome. **A** Heatmap generated by hierarchical clustering of protein levels at 6 months post-HSCT. One patient per row is displayed, with patient characteristics to the left. **B** Comparison of immune reconstitution between the two major clusters from the heatmap. The p values were obtained for clusterxtime interaction terms in linear mixed effect models with time and cluster as additional fixed effects and including a random

At 6 mo after HSCT, complication-prone status was clearly more frequent in cluster 2, where 14/15 patients had complication-prone status vs. 4/10 in cluster 1 (93% vs. 40%, p=0.007 by Fisher’s exact test). Of individual complications, the separation was mainly explained by higher CMV infection incidence in cluster 2 (53% vs. 10%, p=0.04) as well as the major complications of relapse, death, and cGVHD only occurring in cluster 2. Other viral reactivations were distributed evenly in both clusters: HHV-6 reactivation affected 40% of the patients in cluster 2 vs. 20% in cluster 1 (p=0.4 by Fisher’s exact test), EBV and BKV viremias occurred with similar frequencies (13% vs. 30% (p=0.4) and 47% vs. 50% (p=1.0), respectively). The frequencies of aGVHD were similar (67% vs. 60%, p=1.0). The clinically more relevant aGVHD grades 3 and 4 were more common in cluster 2, but the difference was insignificant (23% vs. 10%, p=0.6). Corresponding heatmaps for other timepoints also showed association with major clinical outcomes (supplementary material, Fig. S4).

We next analyzed how these two clusters differed in their overall cellular immune reconstitution parameters. Numbers of T cells (defined as CD3+CD19-), CD4+ T cells (defined as CD3+CD19-CD4+), and B cells (defined as CD3-CD19+) in the circulation were significantly lower from 6-month time point onwards in the complication-enriched cluster 2 when the whole follow-up was included in the statistical analysis (Fig 2B). In cluster 2, total T cells and CD4+ T cells were lower from time point 6 months onwards, B cells during the whole follow-up. Interestingly, CD8 T and NK cell counts did not show differences. We thus can conclude that follow-up-wide immune reconstitution of CD4 T cells and B cells correlated with protein profile status at the 6-month time point (Fig. 2B).

In all, our unsupervised analysis highlighted that CMV reactivation was one of the most defining clinical features for the patients segregated by a specific plasma inflammatory proteome profile. Moreover, this proteome profile was evident prior to failure in specific parts of cellular immune reconstitution.

### Protein coexpression levels are connected to outcome

We analyzed the global evolution of the proteome profile by studying patterns of correlating proteins between our complication-free and complication-prone groups (Fig. 3A-B). Positive and negative correlation coefficients respectively indicate a correlated upregulation or a correlated downregulation of two proteins whereas a correlation coefficient around 0 indicates no coordinated expression of two proteins. The complication-prone patients had a narrow distribution of correlation coefficients peaking at 0 indicating little protein coexpression in early timepoints of 3 and 6 months when compared to complication-free patients. On the contrary, complication-free patients had a positively skewed pattern of correlation coefficients indicating correlated, i.e. coordinated, upregulation of proteins (Fig. 3A). At 12 months after HSCT, at the peak thymopoiesis, the protein correlations of the complication-prone patients become positively skewed while complication-free patients had an even Gaussian distribution of the correlation coefficients, reflecting the homeostasis acquired (Fig. 3A). This is illustrated in the correlation matrix of different correlations between proteins: at the early timepoints in the complication-free patients there is a clear module of coexpressed proteins while in complication-prone patients the protein expression is disorganized. In contrast at the 12-month time point, modules of coexpressed proteins were observed only in the complication-prone patients (Fig. 3B).

**Fig. 3.**
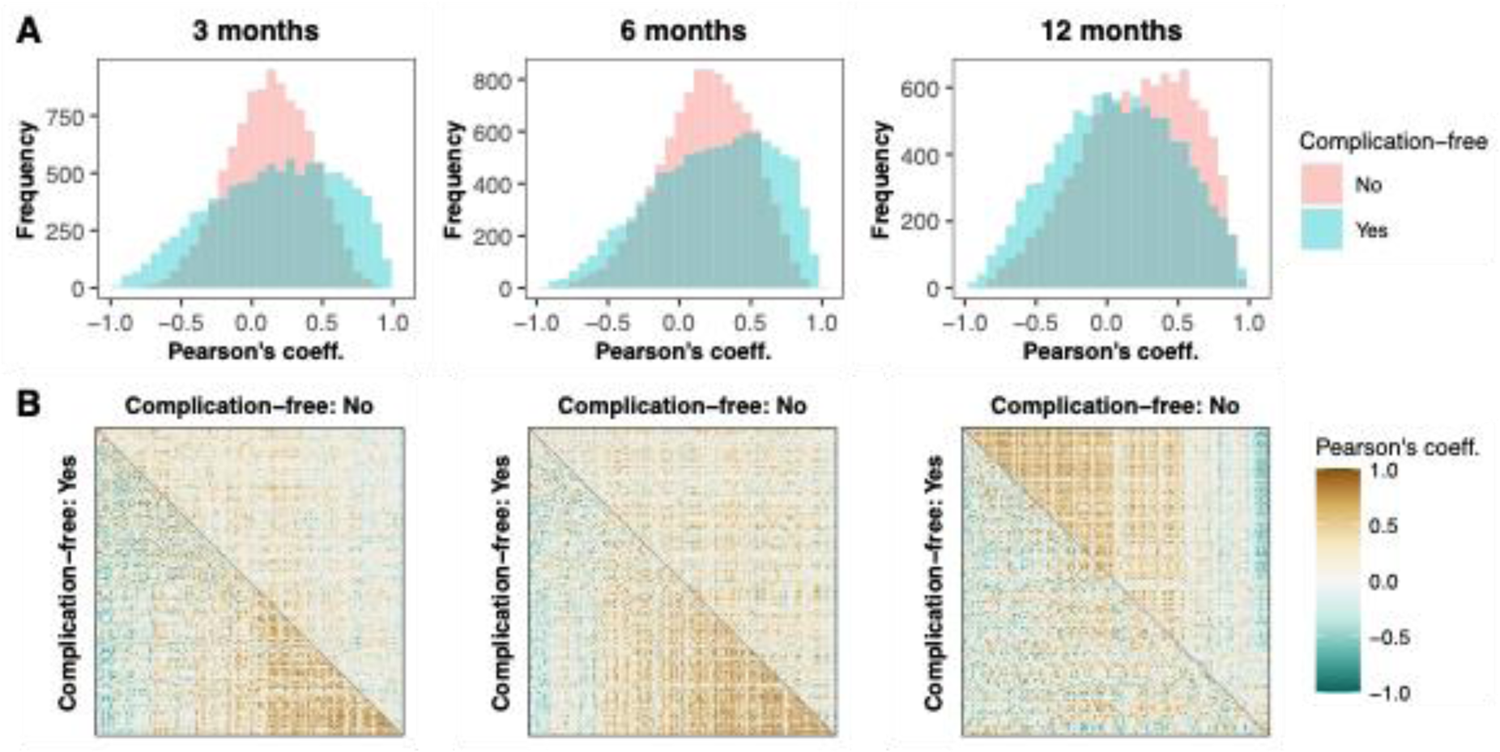
Inter-protein correlations within complication-free and complication-prone groups. A Histograms of Pearson’s correlation coefficients grouped according to complication status B Pearson’s correlation matrices for correlation-free and correlation-prone groups. The proteins were ordered according to the differences in correlation between the two groups by the median strength of correlation differences for each protein.

In summary, the protein correlation patterns over the immune reconstitution period after HSCT are different depending on the outcome. Graphically, the Pearson’s correlation matrices show that patients with more complications have less correlating protein pairs, i.e. a more random protein profile at early time points while the complication-free patients showed more syncronized inflammatory protein kinetics. This was also shown statistically (supplementary material, Table S4), where protein pair correlation level was higher in the complication-free compared to the complication-prone group at early time points. The heterogeneity of the complications included in the complication-prone group most likely explains the disorganized inflammatory protein kinetics. The positively skewed protein correlation pattern in complication-prone individuals one year after HSCT could indicate that achieving immune homeostasis was delayed in the complication-prone individuals.

### Individual protein trajectories are associated with specific outcomes

Our analysis thus far has established that our complication-free and complication-prone patients had qualitatively different proteome profiles over the immune reconstitution after allo-HSCT. Because large-scale proteome analysis is not feasible for use in clinical practice, we continued to analyze specific proteins in relation to specific outcomes over the whole course of our prospective sample set. Here, we applied the maSigPro regression-based analysis which follows a two steps regression strategy to find protein expression values with significant temporal expression changes. We reasoned that the proteins with significantly different expression values over the whole immune reconstitution have the highest potential to be clinically relevant biomarkers. In addition, we tested the protein levels against clinical outcomes at every time point using the Mann-Whitney method in order to identify the clinical correlates for each identified potential biomarker (Fig. 4).

**Fig. 4.**
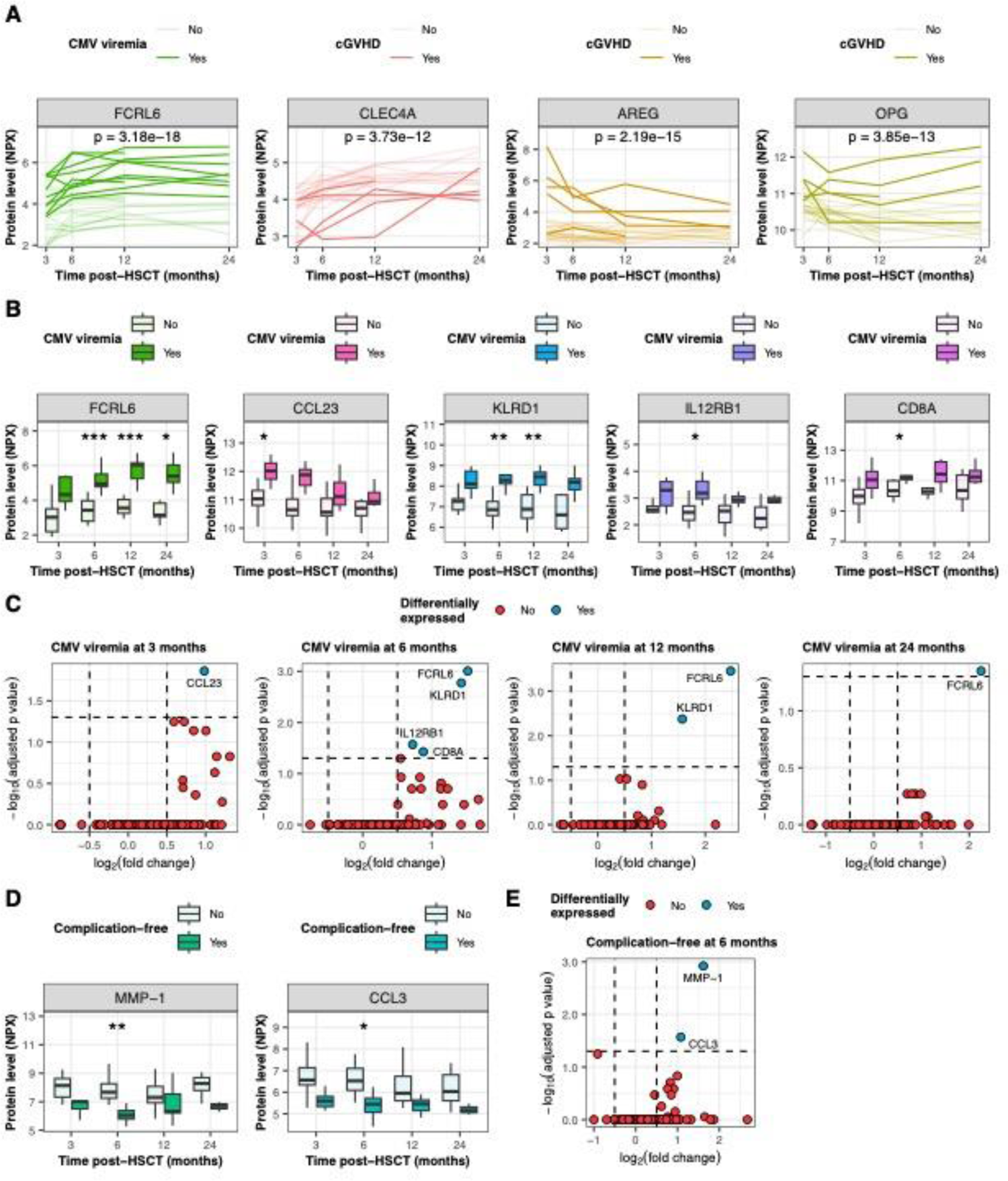
Individual proteins and outcomes. **A** Kinetics of maSigPro-identified significant protein level differences over lime, depending on outcome. One line represents one patient. **B** Timepoint specific comparison of protein levels depending on CMV status. C Levels of differentiation according to outcome and timepoint depending on CMV status. D Timepoint specific comparison of protein levels depending on complication status. E Levels of differentiation according to outcome and timepoint depending on complication status. FCRL6, Fc receptor-like protein 6; CLEC4A, C-type lectin domain family 4 member A; AREG, Amphiregulin; OPG, Osteoprotegerin;; CCL23. C-C motif chemokine ligand 23; KLRD1, Killer cell lectin like receptor D1; IL12RB1, Interleukin 12 receptor subunit beta 1; CD8A, T-cell surface glycoprotein CD8 alpha chain; MMP-1, Matrix metalloproteinase-1 ; CCL3. C-C motif chemokine 3.

Our analysis with maSigPro showed that proteins CLEC4A (C-type lectin domain family 4 member A, also known as DCIR [DC immunoreceptor]), AREG (Amphiregulin), OPG (osteoprotegerin, also known as tumour necrosis factor receptor superfamily member 11B) and FCRL6 (Fc Receptor Like 6) levels showed significantly different expression values over the whole immune reconstitution (Fig. 4A). When we correlated these proteins with clinical outcomes, we found correlations between cGVHD and low CLEC4A (p=3.73 x 10^-12^), high AREG (p=2.19 x 10^-15^) and high OPG (p=3.85 x 10^-13^) (Fig 4A). Moreover, there was a highly significant association between CMV viremia and high FCRL6 (p=3.18 x 10^-18^) (Fig. 4A).

Of the proteins that were significantly different over the whole follow-up period, FCRL6 showed the clearest connection with outcome: for CMV infection at time points 6, 12 and 24 months, log2-fold change was 1.52 (p=9.9 x 10^-4^), 2.47 (p=3.5 x 10^-4^) and 2.25 (p=0.045) (Fig 4B). In time point specific analysis, CMV viremia was also associated with high CCL23, high KLRD1, high IL12RB, and high CD8A, but these correlations were statistically significant in only one or two timepoints and the fold-change differences were lower than with FCRL6 (Fig 4C). For example, higher CCL23 expression levels were significantly associated with CMV reactivation only in the earliest timepoint, after which its expression trajectory was not significantly different from the individuals who did not have CMV reactivation. Only FCRL6 showed significant differences in all time points from the 6-month time point onwards. On the other hand, overall complication-prone status was associated with high MMP-1 and CCL3 (C-C motif chemokine ligand 3, also known as macrophage inflammatory protein 1 alpha, Fig. 4C-E).

### γδ T cells expressing FCRL6 associates with CMV infection

FCRL6 appeared to be the most significant protein differentiating complication-free and complication-prone patients from 6 months after HSCT onwards. Since it was associated with CMV reactivation, we used flow cytometric analysis to further focus on its expression on cytotoxic T cells with known roles to control CMV. Overall, there was a higher trend for FCRL6 expression in CD8+ T cells in patients with CMV viremia than in those without, but the results remained statistically nonsignificant (Fig. 5A). No statistically significant differences of FCRL6 expression between patients with or without CMV viremia were observed in effector-memory, central memory or RA+ effector-memory subsets of CD8+ T cells except for a slight difference at 12-month-time point for central memory subset. We then concentrated on another cytotoxic T cell population, the γδ T cells (defined as CD3+CD8+gamma-delta+), where we observed significantly higher levels of FCRL6 expression in CD8+ γδ T cells on timepoints 6 and 12 months post-HSCT in patients with CMV viremia (Fig. 5B). In addition, the expression of FCRL6 in all γδ T cells was significantly higher in CMV patients at 6 months (Fig. 5B).

**Fig. 5.**
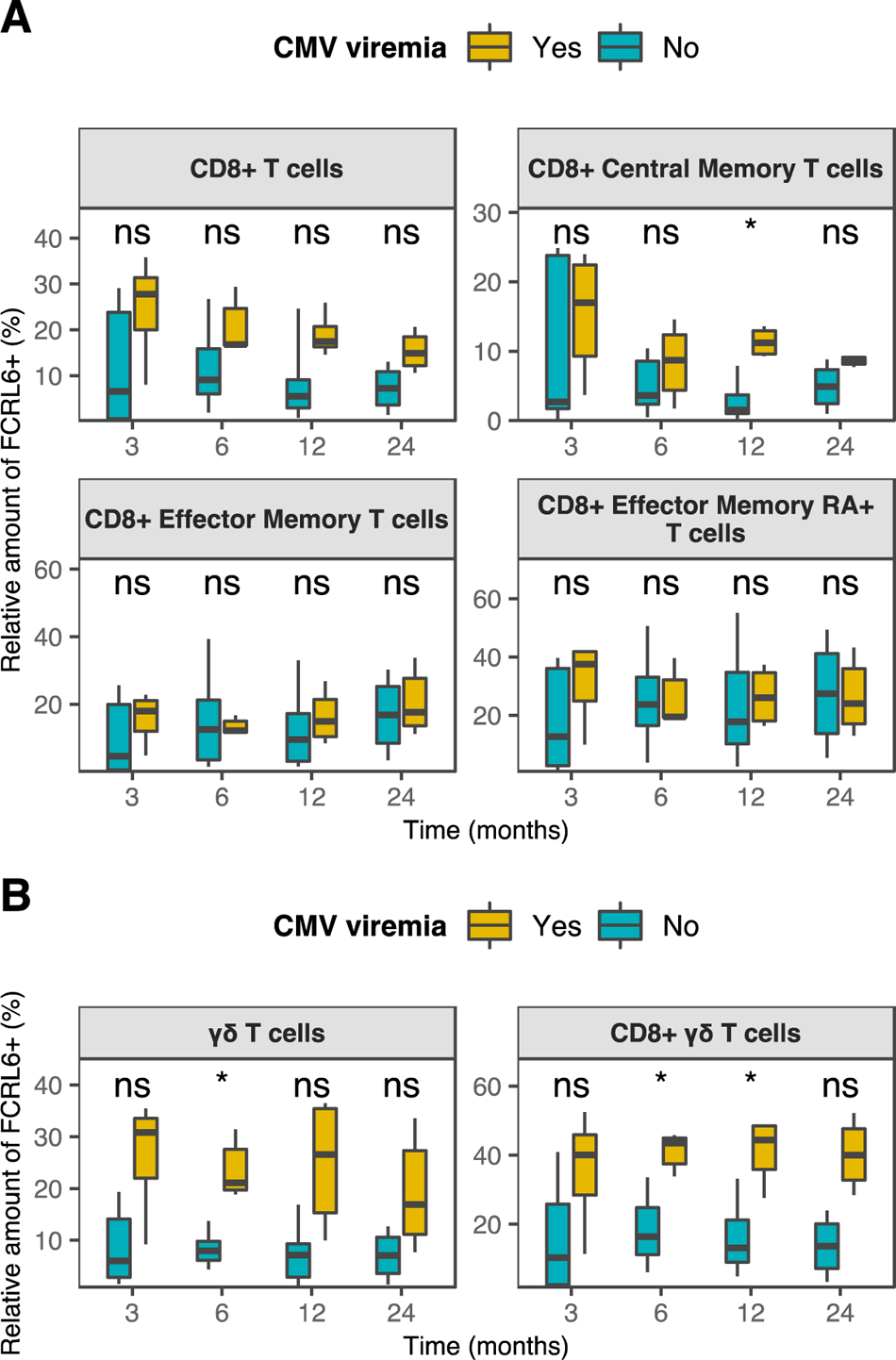
FCRL6-positive cytotoxic T cell subsets in patients with CMV viremia. **A** The box plots show the fraction of FCRL6+ cells of CD8+ T cells, CD8+ central memory T cells, CD8+ effector memory T cells and CD8+ effector memory RA+ T cells at different time points, depending on CMV status. **B** The box plots show the fraction of FCRL6+ cells of all γō T cells and of CD8+ γō T cells, depending on CMV status.

## Discussion

In this single center prospective study of children undergoing allogenic HSCT after myeloablative conditioning as treatment for a hematologic malignancy, we found a clear association between overall plasma protein profile, as well as certain specific proteins, and the complication occurrence.

Hierarchical clustering, an inherently unsupervised process, revealed distinctly different protein profiles with regard to the following outcomes: death, relapse, cGVHD, CMV infection, and immune reconstitution. Even though the predictive value and clinical utility of the method are difficult to establish, hierarchical clustering illustrates the difficulties in denoting specific biomarkers for separate complications since the pathophysiologic processes are greatly intertwined. Delayed immune reconstitution is a well known risk factor for HSCT complications(3). When considering the individual clinical outcomes, the two clusters generated by hierarchical clustering were the most clearly separated by CMV reactivation status, which is also a risk factor for delayed immune reconstitution. After analyzing how different protein expression levels correlated during the immune reconstitution period, we can conclude that the complication-prone individuals showed a clearly divergent pattern of protein correlations, while the good outcome-associated cluster 1 displayed a more synchronized pattern. This chaotic appearance is, by our interpretation, a representation of dysregulated immune reconstitution in individuals that developed complications. These findings were in line with the protein pair analysis, where a good outcome was associated with a clear abundance of positively correlating protein pairs at 3 and 6 months (supplementary material, Table S4).

Among proteins that were higher in CMV-reactivation patients were KLRD1, IL12RB1 (Interleukin 12 receptor subunit beta 1), CD8A and CCL23. All these molecules are associated with cytotoxic responses, i.e. their elevated levels were not surprising in patients experiencing CMV reactivation. The known proinflammatory proteins chemokine CCL3 and metalloproteinase MMP-1 were higher in complication-prone patients, unsurprising since they are known contributors of pathological inflammatory processes(20), (^21^), (^22^), (^23^). AREG, an autocrine growth factor and a mitogen for astrocytes, Schwann cells and fibroblasts(24), was higher in patients with cGVHD at 3 months post-HSCT, probably reflecting the verified relevance of AREG in aGVHD(25) and late aGVHD(26). Other cGVHD-associations were higher levels of OPG (Osteoprotegerin or TNF receptor superfamily member 11b) and lower levels of CLEC4A.

The most noticeable change between patients with CMV viremia and those without was the protein levels of FCRL6 (Fc receptor-like 6, Fig. 4), an immunoregulatory receptor whose expression is restricted to cytotoxic lymphocytes(27,28). The mechanism of action of FCRL6 is largely unknown, but its interaction with HLA-DR molecules led to reduced cytotoxicity of NK cells towards tumor cells similarly to LAG-3, which is a known marker for effector cell exhaustion(29,30). Hence, FCRL6 has been suggested to be a possibly druggable check-point molecule to reverse the effector cell exhaustion. It could also serve as a potential biomarker for relapses(29). To our knowledge, no association between FCRL6 expression and CMV infection has been shown before. While FCRL6 has been shown to be upregulated in chronic inflammatory states(31), we found no connection with e.g. cGVHD or other viral infections. The difference in FCRL6 levels was not explained by standard lymphocyte cell counts, since its expression is restricted to cytotoxic CD8+ and NK cells which showed comparable immune reconstitution between clusters 1 and 2 at the 6-month time point. Since the FCRL6 has no known soluble function, its measurable plasma levels are likely due to increased receptor shedding from FCRL6 positive cells.

Fraction of FCRL6 expressing CD8+ γδ cells was consistently higher in patients that had CMV reactivation (Fig. 5). γδ T cells have a central role in defense against CMV reactivation post-HSCT, and higher levels are associated with higher survival and lower relapse rates(32). The important role of CMV control by γδ T cells has also been established in the αβ T cell-depleted haploidentical HSCT setting(33). However, FCRL6 expressing γδ T cells have not been studied before. Recent findings indicate that γδ T cells can control CMV not through direct CMV-antigen recognition but instead by sensing the upregulated HLA molecules during the CMV infection(34). One receptor involved in this sensing could be FCRL6 whose known ligand is HLA-DR. Our results thus highlight the role of γδ T cells in controlling CMV and reveal a new receptor potentially involved in activation or exhaustion of γδ T cells. γδ cells do not cause GvHD so they offer an interesting cellular source for adoptive cellular therapies to treat CMV reactivation(35). Future studies should focus on understanding the functional significance of FCRL6 on γδ T cells.

The relatively small sample size of our cohort did not allow for division into training and validation groups, but our setup with multiple time point measurements in addition to great sample cover, make the data robust. Ideally, time points would have been more numerous and closer to each other, yielding yet greater information on sequence of events as well as more power to time-dependent analyses. Even though not a weakness per se, our multiple parameter data was under significant strain of multiple testing correction, which might have masked some meaningful true associations.

Proteomics screening methods offer a powerful tool to reveal new biomarkers or targets for novel therapies. In our pediatric cohort, the proteomic approach revealed several potential biomarkers that showed different kinetics in patients with and without complications post-HSCT. Although not directly clinically translatable, plasma protein profiles seem to tell us more than levels of individual proteins alone. Moreover, the inhibitory FCRL6 receptor emerged as a potentially relevant biomarker or a therapy target on cytotoxic T cells for CMV reactivation.

## Data Availability

All data produced in the present study are available upon reasonable request to the authors

## Acknowledgements

We thank research nurse Pia Valle, and lab technicians Marjo Rissanen, Tamas Bazsinka and Turku University for indispensable assistance.

